# Genome-wide association study of human pineal gland volume as proxy for melatonin secretion

**DOI:** 10.1101/2025.03.10.25323654

**Authors:** Peng Xu, Mohammed Aslam Imtiaz, Daniel Rusman, Santiago Estrada, Martin Reuter, Monique M.B. Breteler, N. Ahmad Aziz

## Abstract

The pineal gland is the primary source of melatonin synthesis and secretion. Reduced pineal gland volume results in lower melatonin levels and has been linked to sleep impairment, as well as many (age-related) metabolic and neurodegenerative disorders. However, its genetic architecture remains unknown. Using brain imaging and genotype data from the UK Biobank (N=38,254) and Rhineland Study (N=5,286), we conducted the first genome-wide association study (GWAS) of pineal gland volume, demonstrating 19% heritability and identifying 34 genome-wide significant loci, with highly concordant results between the two independent cohorts. Gene mapping and functional annotation highlighted multiple genes and pathways involved in numerous traits including sleep, metabolism, neurodevelopment and neurodegeneration. Mendelian randomization indicated a causal relationship between pineal gland volume and daytime napping. Our findings elucidate the genetic architecture of pineal gland volume and identify candidate genes and molecular pathways that may affect sleep/circadian rhythm, metabolism, neuronal development and brain health.

## Introduction

The pineal gland (also known as the “epiphysis”), a key neuroendocrine structure located in the epithalamus near the center of the brain, is the main source of melatonin synthesis and secretion in humans^1^. Reduced pineal gland volume has been associated with both aging^2^, and many different diseases, including sleep disorders^3^, diabetes^4^, cardiovascular diseases^5^, psychosis/schizophrenia and Alzheimer’s disease^6,7^. These associations are largely attributed to melatonin’s important role in the regulation of various physiological processes such as circadian rhythms^8^, glucose and insulin signalling^9^, as well as its antioxidant, immunomodulatory and neuroprotective properties^10,11,12^. Given that pineal gland volume is strongly related to the total amount of daily melatonin secretion^13,14^, exploring the genetic architecture of pineal gland volume could provide novel insights into the genetic determinants of melatonin synthesis and secretion, with important implications for the understanding of a broad range of physiological and disease mechanisms.

Prior twin- and family-based studies have indicated that genetic factors could underlie interindividual differences in melatonin production^15,16^. However, a previous small-scale genome-wide association study (GWAS) in a Taiwanese cohort of 2373 individuals, which used urinary 6-hydroxymelatonin sulfate-to-creatinine ratio as the outcome, failed to identify any genome-wide significant loci^17^. Due to its strong circadian rhythmicity, accurate quantification of melatonin production requires sampling at specific timepoints throughout the day, whether in plasma, saliva or urine^18^. This complicates direct measurement of melatonin production in biofluids in large-scale population-based studies, which are necessary for sufficiently powered GWAS studies. Moreover, it is also unlikely that melatonin secretion during a single day/night cycle would accurately reflect its long-term rate of production. Consequently, the genetic basis of melatonin production has remained elusive.

To address the abovementioned challenges, recently we developed a fully automatic, multi-modal parcellation procedure for volumetric segmentation of the pineal gland on brain magnetic resonance images (MRI), enabling accurate, high-throughput quantification of pineal gland volume in large-scale population-based studies^19^. Leveraging this tool, here we performed the first-ever GWAS on pineal gland volume, as a proxy for melatonin production, in the UK Biobank Study, followed by replication of the findings in the population-based Rhineland Study, as well as a range of *in silico* functional analyses to contextualize the biological relevance of our findings. Importantly, in our meta-GWAS we discovered 34 genome-wide significant loci associated with pineal gland volume, which could be linked to a broad range of traits, including those related to sleep, neurodevelopment and neurodegeneration. Notably, two-sample mendelian randomization (MR) yielded evidence for a causal relationship between pineal gland volume and daytime napping.

## Methods

### Participants

We included 38254 participants from the UK Biobank Study in the discovery GWAS following quality control of both imaging and genotyping data (**Supplementary Figure 1**). Applying similar quality control procedures, 5286 individuals from the Rhineland Study were included in the replication GWAS (**Supplementary Figure 2**). We also conducted a meta-GWAS by combining the discovery and replication GWAS summary statistics (N = 43540). Approval for conducting the UK Biobank Study was received from the National Information Governance Board for Health and Social Care and the National Health Service North West Centre for Research Ethics Committee (Ref: 11/NW/0382). The Rhineland Study protocol was approved by the ethics committee of the University of Bonn Medical Faculty (Ref: 338/15), and the study was conducted according to the International Conference on Harmonization Good Clinical Practice standards (ICH-GCP). All participants in both cohorts provided written informed consent in accordance with the Declaration of Helsinki.

### Imaging data

In the UK Biobank Study, imaging data were acquired using Siemens Skyra 3T MRI scanners equipped with 32-channel head coils. Initially, a single scanner located in Cheadle Manchester, was designated for the UK Biobank Study, later expanding to include two additional identical scanners in Newcastle and Reading^20^. The full protocols and MRI sequence parameters in the UK Biobank Study can be found in the online documentation (<u>biobank.ctsu.ox.ac.uk/crystal/crystal/docs/brain_mri.pdf</u>). In the Rhineland Study, imaging data were collected using two identical 3T Siemens MAGNETOM Prisma MRI scanners, each equipped with 64-channel head-neck coils, at two examination centers in Bonn, Germany^21^.

For segmentation of the pineal gland we applied our automated deep learning-based method ‘FastSurfer-HypVINN’^19^ on 0.8 mm isotropic T1- and T2-weighted images from the Rhineland Study, as well as 1.0 isotropic T1-weighted images from the UK Biobank Study. Importantly, we had previously validated the accuracy of ‘FastSurfer-HypVINN’ against manually segmented ground-truth MR images from both the Rhineland Study and UK Biobank Study. Furthermore, estimated total intracranial volume (ETIV) for both cohorts was derived using the FreeSurfer (version 6.0) standard processing pipeline^22,23^.

### Genetic data

A standard protocol was followed for the quality control of genetic data in the UK Biobank Study^24^. Briefly, single nucleotide polymorphisms (SNPs) were filtered out based on call rate (<95%), deviation from Hardy-Weinberg equilibrium (HWE) (p-value <1e-6) and minor allele frequency (MAF) <1%. The genetic samples were checked for call rate, ethnic outliers, samples with abnormal heterozygosity and filtered accordingly. The genetic data in the UK Biobank Study were imputed using the HRC reference panel. SNPs with an imputation score <0.3 were excluded. In the Rhineland Study, genotyping was performed using Omni 2.5 Exome array. Genotypes from signal intensity files were called using Genome Studio (version 2.0.5). Quality control was performed separately for SNPs and samples using PLINK (version 1.9). SNPs were excluded based on missing rate (>98%), deviations from HWE (p-value <1e-6), minor allele frequency (MAF <1%), and if palindromic with MAF >0.4, whereas samples were filtered based on call rate (>95%), abnormal heterozygosity, and ethnic outliers deviating from European ancestry estimated using EIGENSTRAT (version 16000)^25^. In the Rhineland Study, after quality control, genetic data were imputed using Impute2^26^ software with 1000 genome version 3 phase 5 reference panel (https://www.nature.com/articles/nature15393). SNPs with an imputation score <0.3 were excluded^27^.

### Discovery, replication and meta-GWAS analysis

Regenie^28^ was used to conduct the GWAS analyses, while accounting for cryptic relatedness through application of linear mixed-effects models. As outcome we used the residuals from the model of the pineal gland volume as a function of age, age-squared, sex, ETIV, and the first 10 genetic principal components, and only in the UK Biobank Study, additionally of MRI scanner site, batch, and array (Model 1). Before GWAS analysis, all phenotypic variables were transformed into a normal distribution using inverse-rank transformation. In a second model (Model 2), we further adjusted for the volumes of the frontal, parietal, temporal and occipital cortices to account for pineal gland volume reduction due to non-specific generalized brain atrophy. For the meta-GWAS analysis, we used METAL^29^ to run a fixed effects inverse variance-weighted meta-analysis using the GWAS summary statistics of Model 1 and 2 from both cohorts. In a sensitivity analysis, we also included total brain volume as an extra covariate to Model 2 to assess whether the discovered genetic associations with pineal gland volume were independent of generalized brain atrophy.

### Risk loci definition

We used the Functional Mapping and Analysis of GWAS (FUMA) platform to identify genomic risk loci^30^. Genome-wide significant SNPs in relatively high linkage disequilibrium (LD) (i.e., r^2^ ≥ 0.6) with nearby SNPs were used to define genomic risk loci, merging LD blocks of independently significant SNPs within 250 kb of each other into a single genomic locus. Within each genomic locus, we defined the *lead* SNPs as those SNPs that are independent of one another at r^2^ < 0.1, using the 1000 Genome Phase 3 version 5 reference panel.

### Gene mapping

Positional mapping of SNPs to genes (physical distance within 10 kb) was performed using ANNOVAR^31^. Moreover, SNPs were also mapped to genes based on expression quantitative trait loci (eQTL) and chromatin interaction mapping. For eQTL mapping, we specifically selected brain tissue expression data from GTEx v8^32^, BRAINEAC^33^, PsychENCODE^34^, and Common Mind Consortium^35^. FUMA performs three-dimensional DNA-DNA chromatin interaction mapping, where genes are identified based on significant chromatin interactions between pineal gland volume associated regions and nearby and distant genes in PsychENCODE EP links^33^, PsychENCODE Promoter anchored loops^35^, HiC data from brain tissues, including adult cortex^36^, fetal cortex^36^, dorsolateral prefrontal cortex^37^, hippocampus^37^, HiC data from neural progenitor cell lines^37^, as well as genes overlapped with a predicted enhancer region (250 bp up- and 500 bp downstream of transcription start sites) available in 17 brain-related repositories from the Roadmap Epigenomics Project^38^ (i.e., E053, E054, E067, E068, E069, E070, E071, E072, E073, E074, E081, E082, E003, E008, E007, E009, E010, E057). The significant 3D chromatin interactions were identified using the default FDR threshold P-value < 1e-6, and visualized using circus plots.

### Functional mapping

Using the GENE2FUNC function in FUMA, the biological context of the mapped genes was further investigated by assessing their enrichment in 53 GTEx tissue-specific gene expression gene sets^32^, curated MSigDB gene sets^39^, and GWAS Catalog database of reported genetic-variant trait associations (e110_r2023-07-20)^40^. We used the default Bonferroni adjusted P-value < 0.05 for inferring statistical significance, and required that at least two input genes overlapped with a tested gene set.

### Genome-wide gene-based association study and gene-set enrichment analyses

MAGMA^41^ was used to run the gene-based analyses using the meta-analysis summary statistics of pineal gland volume as input. For gene-set analyses, 17,023 curated gene sets available in MSigDB version 2023.1.Hs^42,43^ were used to test whether gene-based P-values among all 18,883 protein-coding genes were lower for those genes within pre-defined gene sets that represent biological processes and disease pathways compared to other genes.

### SNP heritability and genetic correlation

SNP heritability was estimated through LD Score regression (LDSC)^44^ using GWAS summary statistics. LDSC was also used to estimate the genetic correlation of pineal gland volume with sleep related traits (i.e., chronotype^45^, daytime napping^46^ and sleep duration^46^), neurological traits (i.e., cognition^47^), neurodegenerative diseases (i.e., Alzheimer’s disease^48^ and Parkinson’s disease^49^), as well as 101 brain-related regional volumetric imaging-derived endophenotypes^50^. Chronotype was assessed with a shortened version of the Morningness-Eveningness Questionnaire^45^. Daytime napping was assessed through self-reports (i.e., based on answers to the question “Do you have a nap during the day?”), while sleep duration was derived from accelerometry data^46^. Cognition was defined as a latent factor underlying multidimensional cognitive functioning / intelligence^47^. The 101 regional volumetric imaging-derived endophenotypes were generated as described previously^50^ (an overview is provided in **Supplementary Table 18**). Genetic correlation was calculated using HapMap3 SNPs only, while the Benjamini-Hochberg FDR procedure was applied to correct for multiple testing.

### Two-sample mendelian randomization

We employed a two-sample MR approach, as implemented in the R package Twosample MR (v 0.5.7), to test whether pineal gland volume was causally associated with sleep-related and/or neurological/neurodegenerative traits. The inverse variance weighted regression method (IVW)^51^ was used as the primary test, followed by sensitivity analyses using other methods robust to instrumental variable bias due to horizontal pleiotropy, including MR-Egger regression^52^, weighted median and mode-based methods^53^. All P-values obtained were adjusted for multiple comparisons using FDR correction. Heterogeneity in the IVW and MR-Egger regression estimates was assessed using Cochran’s Q-test^54^. In the presence of heterogeneity, radial MR^55^ was used to detect outliers in instrumental variables. Moreover, using the same approach, we also tested for reverse causation. The presence of weak instrument bias^56^ was assessed using the F-statistic^57^, with values <10 indicating weak genetic instruments.

## Results

### Population characteristics

The mean age of the study participants was 64.4 ± 7.7 years (52.7 % women) in the UK Biobank Study, and 55.6 ± 13.6 years (57.3 % women) in the Rhineland Study. The mean volume of the pineal gland was 209 ± 78 mm^3^ in the UK Biobank Study and 229 ± 78 mm^3^ in the Rhineland Study participants (**Supplementary Table 1**).

### GWAS analysis

In total, 7,308,990 and 7,303,002 SNPs were included in the discovery and the replication GWAS, respectively. In the discovery GWAS, we found 3860 genome-wide significant SNPs in Model 1, including 145 independent significant SNPs and 53 lead SNPs, comprising 37 genomic risk loci (**Supplementary Figure 3a,d** and **Supplementary Tables 2-4**). These results remained largely similar in Model 2, which additionally accounted for generalized cortical atrophy, with 3858 SNPs achieving genome-wide significance, including 145 independent significant SNPs and 53 lead SNPs, mapping to 36 genomic loci (**Figure 1a**,**d** and **Supplementary Tables 5-7**). In the replication GWAS, about two-thirds of these loci were replicated (Model 1: **Supplementary Figure 3b,e** and **Supplementary Table 2**; Model 2: **Figure 1b**,**e** and **Supplementary Table 5**). Specifically, with the replication threshold defined as an FDR-corrected P-value < 0.05 in Model 2, 22 genomic loci were replicated (**Table 1**, **Supplementary Tables 5-7**). Notably, rs1892277 on chromosome 6 was the strongest hit in the discovery GWAS (P = 1.90 × 10^-72^). However, this SNP was not included in the replication GWAS due to quality control criteria, because it was palindromic with a MAF > 0.4. Nevertheless, the tagged genomic locus was replicated using the proxy SNP rs10948679, which is in strong LD (r^2^ = 0.60 in European ancestry) with rs1892277 (P_FDR_ = 9.55 × 10^-11^). Importantly, rs10948679 was also the strongest signal in the replication GWAS, further validating this novel association.

**Figure 1:**
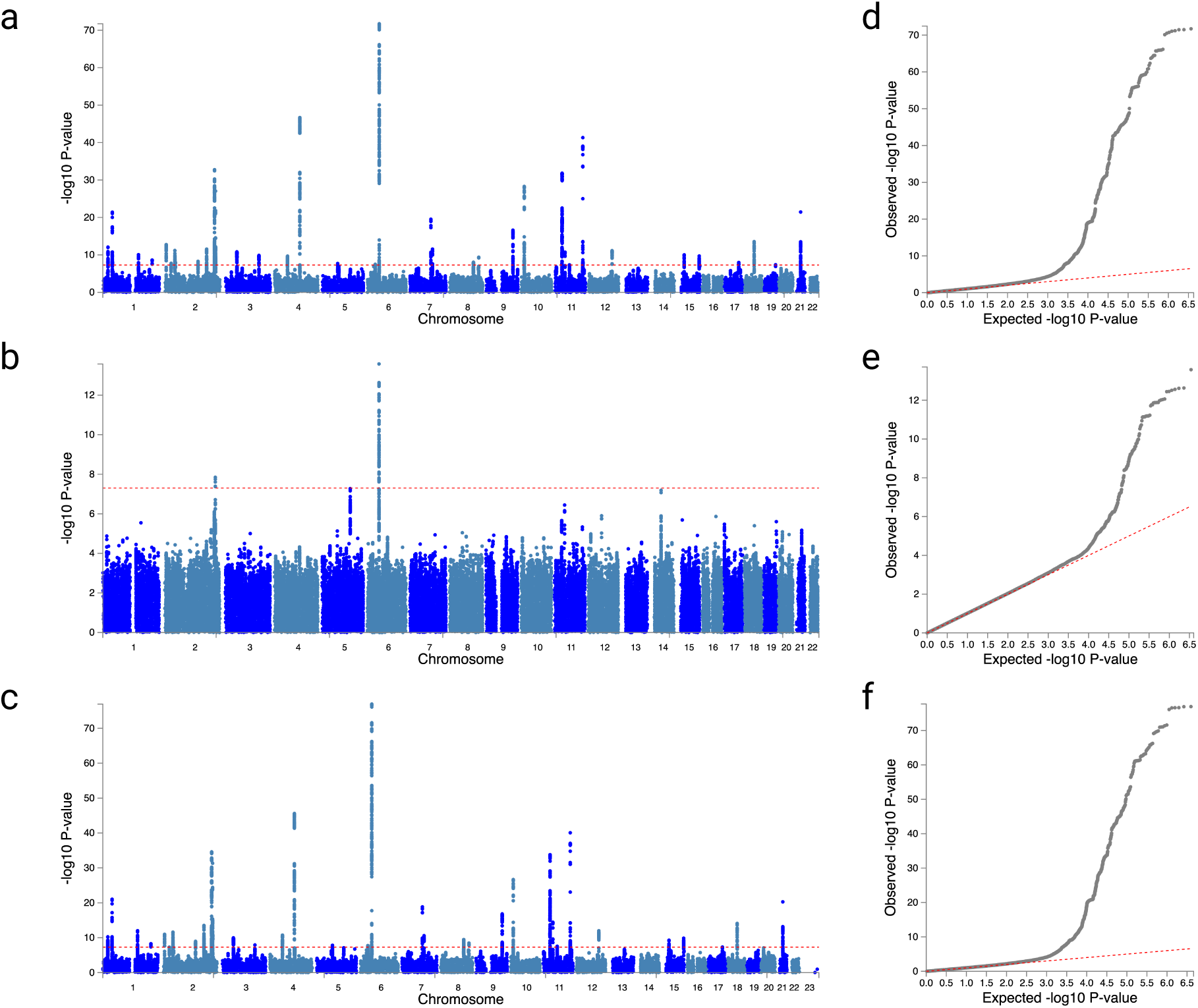
Genomic loci associated with pineal gland volume. Manhattan plots illustrating the −log_10_(P) statistic obtained from Model 2 for the **a.** discovery GWAS, **b.** replication GWAS, and **c.** meta-GWAS. The horizontal red dashed lines indicate the threshold for genome-wide significance (p < 5 x 10^-8^). QQ plots displaying observed versus expected −log_10_(P) statistic from the **d.** discovery GWAS, **e.** replication GWAS, and **f.** meta-GWAS. Red dashed lines represent the −log_10_(P) threshold associated with the null hypothesis of no association. **Abbreviation:** GWAS, genome-wide association study; QQ plot, quantile-quantile plot.

**Table 1.**
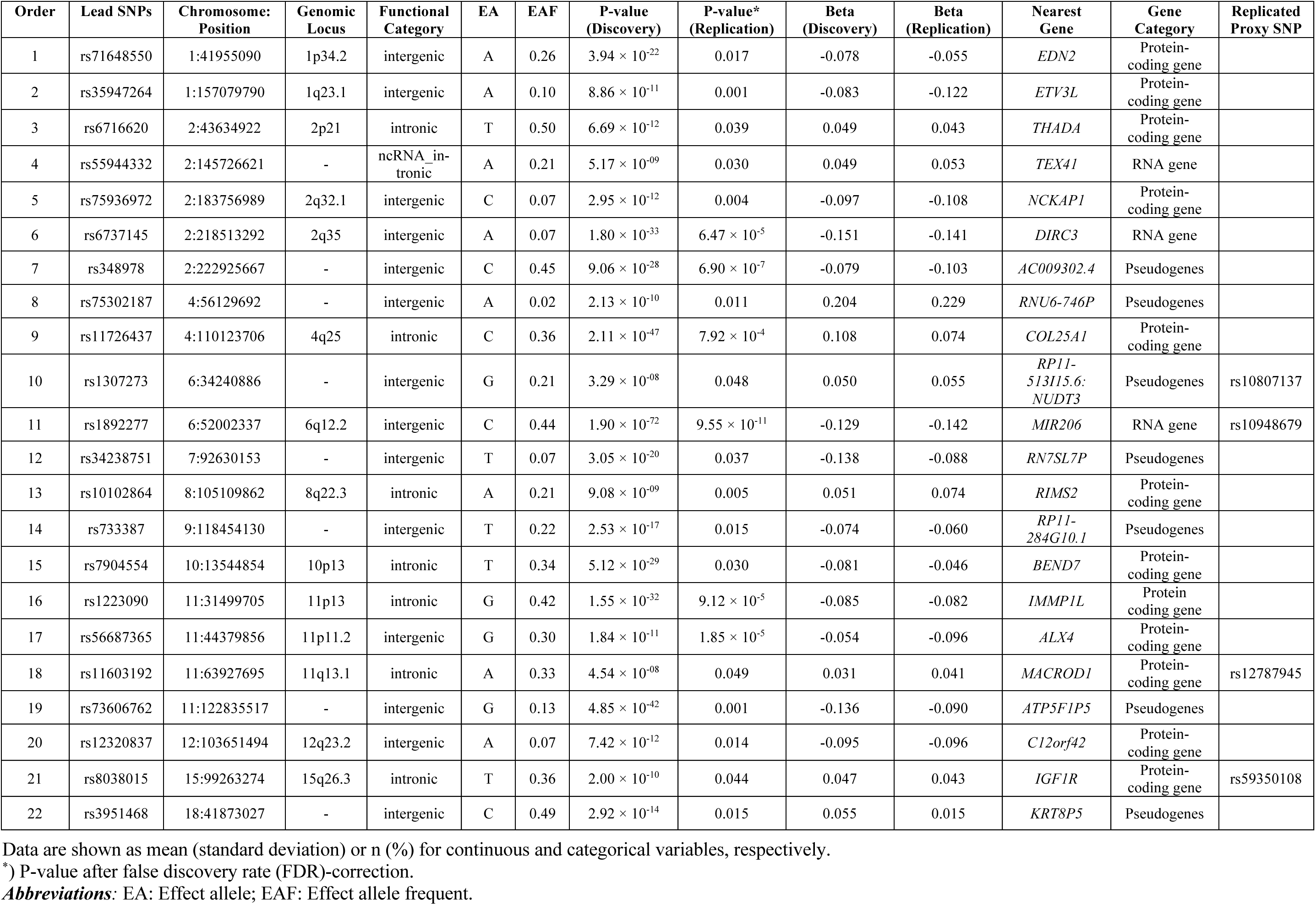
Genomic loci associated with pineal gland volume overlapping in both discovery and replication GWAS.

To identify the most robust signals, we further conducted a meta-GWAS by combining the UK Biobank Study and Rhineland Study datasets, including a total of 8,017,385 unique SNPs. In Model 1, we detected 4041 candidate SNPs, 148 independent significant SNPs and 53 lead SNPs, spanning 35 genomic loci (**Supplementary Figure 3c,f** and **Supplementary Tables 8-10**). Similarly, in Model 2, we identified 3819 candidate SNPs, 141 independent significant SNPs and 51 lead SNPs, comprising 34 genomic loci (**Figure 1c**,**f** and **Supplementary Tables 11-13**). The sizes of the genomic loci ranged from 9.8 kb to 1434.1 kb (**Figure 2a**). Using Model 2 results of the meta-GWAS, SNP-based heritability (*h*^2^) for pineal gland volume was estimated at 0.190 ± 0.024, indicating that approximately 19% of the variability in pineal gland volume can be attributed to common genetic variants.

**Figure 2:**
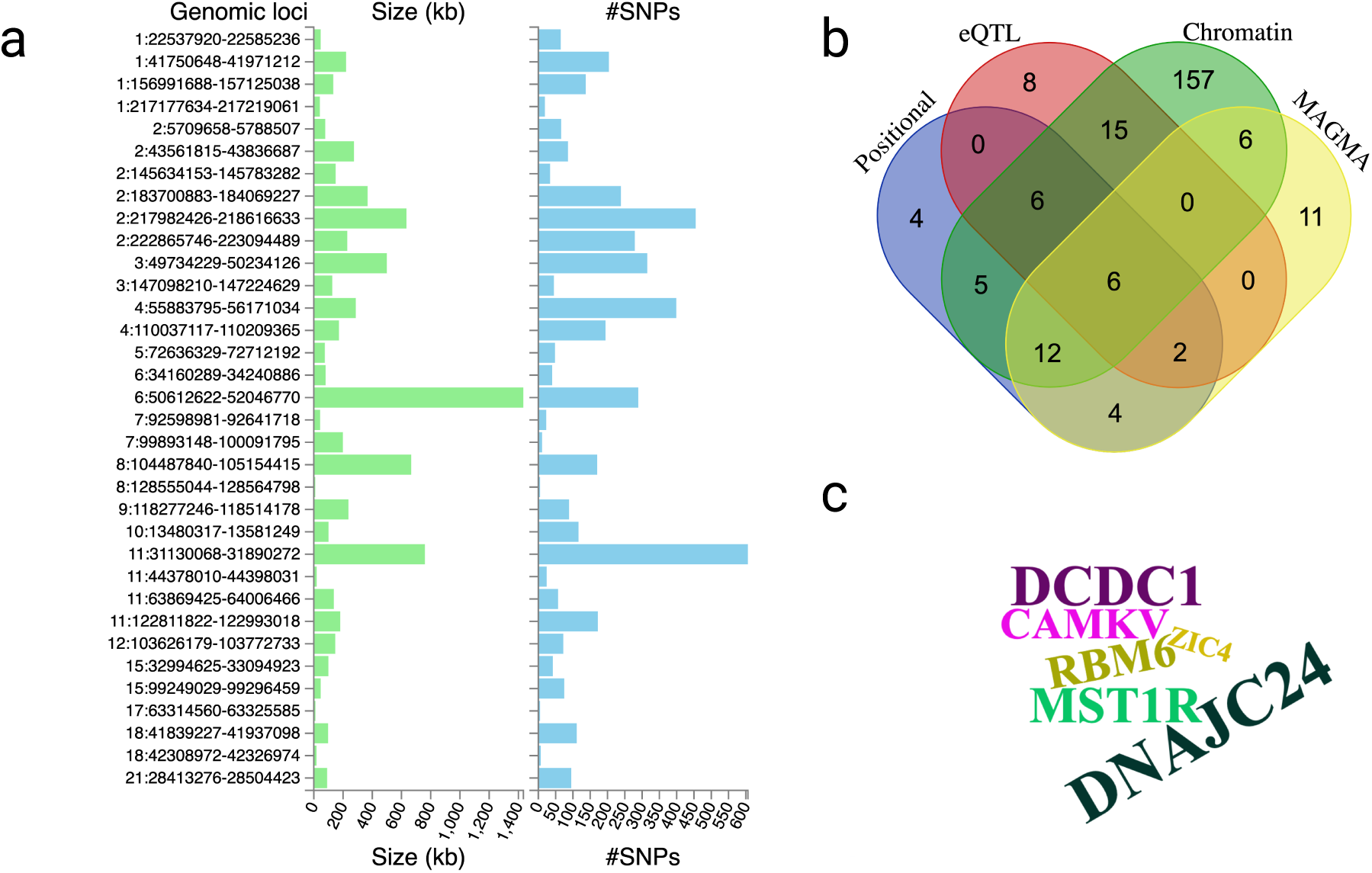
Gene mapping and functional annotation of 34 loci associated with pineal gland volume in the meta-GWAS. **a.** Overview of genomic loci sizes and distribution of candidate SNPs. **b.** Venn diagram depicting the number of genes mapped by the four different strategies. **c.** Word cloud plot displaying the six genes identified through all four strategies, with the size of the word representing –log_10_(P) values from the meta-GWAS. **Abbreviation:** eQTL, expression quantitative trait loci; GWAS, genome-wide association study; MAGMA, multi-marker analysis of genomic annotation; SNP, single nucleotide polymorphism.

To further assess inflation from potential confounding factors, we used LDSC. In the discovery GWAS, the LDSC intercept of pineal gland volume was 1.109 (Model 1) and 1.110 (Model 2), while in the replication GWAS, it was 1.029 (Model1) and 1.026 (Model 2). For the meta-GWAS, the LDSC intercept was 1.030 in both Model 1 and 2, indicating well-controlled population stratification and minimal other confounding effects. Therefore, we used the meta-GWAS results of the fully adjusted model (i.e., Model 2) for further downstream analyses.

Our sensitivity analysis, in which we also adjusted for total brain volume in addition to all the covariates included in Model 2, yielded the same genomic loci as in Model 2 (**Supplementary Table 14 and Supplementary Figure 4**), indicating that the genetic associations with pineal gland volume are independent of generalized brain atrophy.

### Gene mapping and functional annotation

For each of the 22 genomic loci that reached statistical significance in both the discovery and replication GWAS, we identified the nearest genes based on their physical proximity to the lead SNPs in the discovery GWAS, comprising of 12 protein-coding genes, three RNA genes and seven pseudogenes (**Table 1**). Moreover, we functionally annotated all candidate SNPs (n = 3819), defined as those in LD (r² ≥ 0.6) with any of the independent significant SNPs within each genomic locus, identified in the meta-GWAS Model 2 (**Supplementary Table 11**).

Gene-based mapping through multi-marker analysis of genomic annotation (MAGMA), resulted in the discovery of 41 unique genes (**Supplementary Figure 5** and **Supplementary Table 15**). In addition, positional, expression quantitative trait loci (eQTL) and chromatin-interaction mapping led to the identification of 225 unique genes (**Supplementary Table 16**). Notably, six protein-coding genes were consistently highlighted by all four gene-mapping strategies (**Figure 2b**,**c**), indicating robust effects on pineal gland volume. Moreover, through MAGMA-based gene-set analysis we identified the gene-set *GOMF_INSULIN_BINDING* to be significantly related to pineal gland volume (Bonferroni corrected P-value = 3.99 × 10^-3^), paralleling previous findings suggesting a role of the pineal gland and melatonin in diabetes and insulin signaling^4,9^.

For 572 out of our 3819 candidate SNPs, associations with multiple other traits and phenotypes were also reported in previous studies enlisted in the GWAS catalog, including insomnia, blood pressure, height and body composition (**Supplementary Figure 6** and **Supplementary Table 17)**. For genes mapped through positional, eQTL and chromatin-interaction mapping, we also performed a positional gene-set enrichment analysis that indicated enrichment of these genes in 13 specific genomic regions (**Supplementary Figure 7**). Additionally, using the GWAS catalog database, we identified related traits associated with these gene sets, including brain morphology, sleep duration and intelligence (**Supplementary Figure 8**).

### Genetic correlations with brain structures, sleep traits and neurodegenerative disorders

We performed genetic correlation analyses between pineal gland volume and 101 brain imaging-derived volumetric endophenotypes, revealing strong correlations with several of these outcomes (**Figure 3**, **Supplementary Table 18**). Specifically, cerebrospinal fluid (CSF) volume (r_g_ = 0.32, P_FDR_ = 1.37 × 10^-6^), and volumes of the cerebellar vermis lobules I-V (r_g_ = 0.20, P_FDR_ = 0.025) and the right isthmus cingulate (r_g_ = 0.22, P_FDR_ = 0.034) demonstrated significant genetic correlations with pineal gland volume.

**Figure 3:**
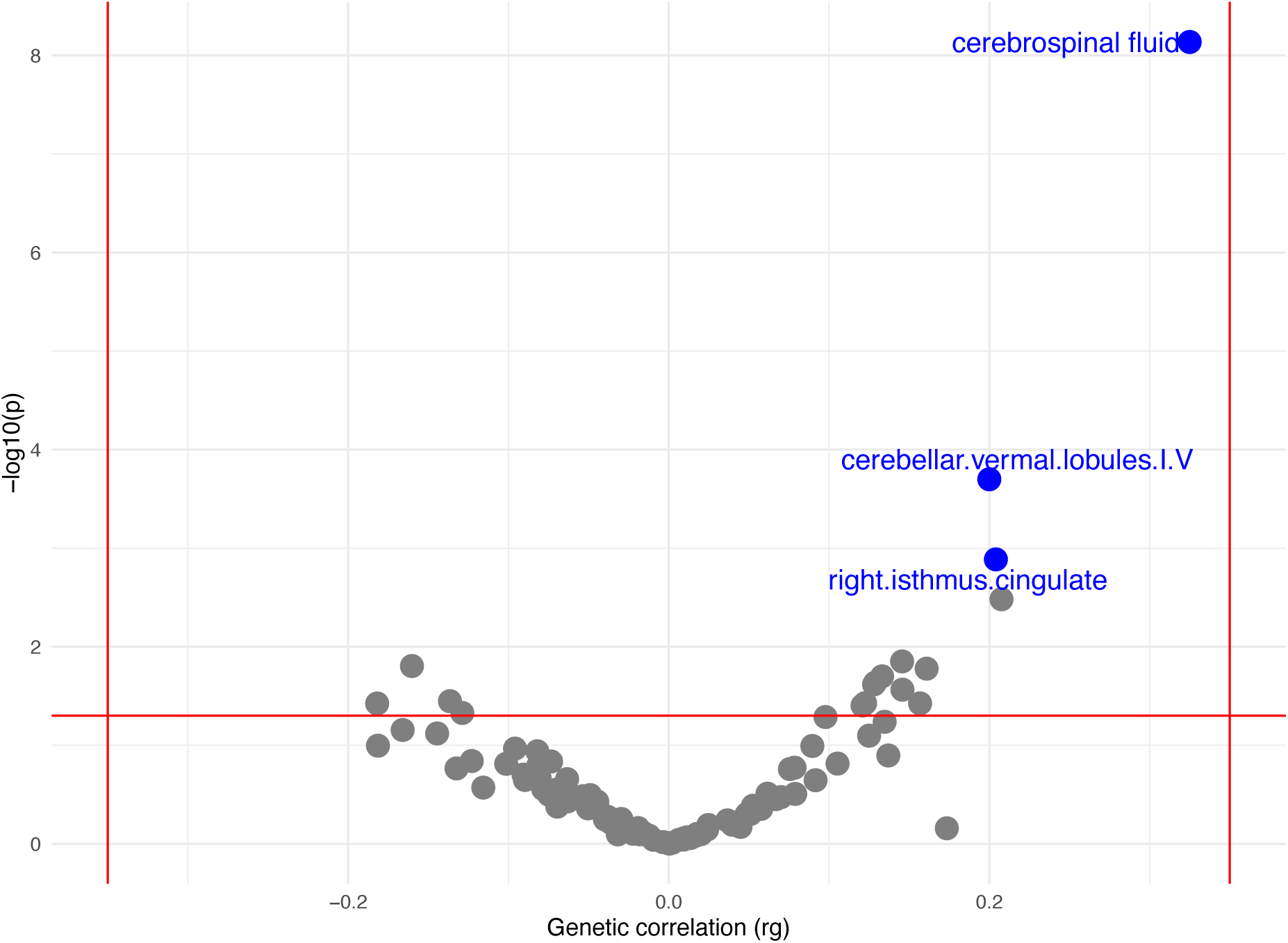
Genetic correlations between pineal gland volume and volumes of 101 brain imaging-derived endophenotypes. Blue and grey dots represent statistically significant (after FDR-correction) and non-significant correlations, respectively. Regions surviving FDR-correction are tagged. The red horizontal line represents the threshold for nominal statistical significance (i.e., p = 0.05). **Abbreviation:** CSF, cerebrospinal fluid; FDR, false discovery rate; GWAS, genome-wide association study; rg, genetic correlation coefficient.

We found non-significant trends for associations of pineal gland volume with daytime napping (r_g_ = −0.06, P = 0.053), chronotype (r_g_ = −0.217, P = 0.076), sleep duration (r_g_ = 0.051, P = 0.099), and Alzheimer’s disease (r_g_ = −0.185, P = 0.069) (**Supplementary Table 19**).

### Mendelian randomization for sleep-related and neurological traits and disorders in relation to pineal gland volume

We performed bidirectional MR studies to explore potentially causal relationships between pineal gland volume, sleep-related and neurological traits and disorders. As sleep-associated traits, we focused on daytime napping, chronotype and sleep duration, all of which have been linked to melatonin levels in previous studies. As neurological traits and disorders, we included cognition and the two most common age-associated neurodegenerative diseases, i.e., Alzheimer’s disease and Parkinson’s disease, which have also been associated with changes in melatonin levels.

Using 31 genetic instruments, in forward MR we discovered a suggestive significant association between pineal gland volume and daytime napping (**Supplementary Table 20**). However, the Cochran’s Q statistic indicated the presence of significant heterogeneity in both IVW and MR-Egger estimates (P < 3.1 × 10^-7^). To address this, we applied radial MR to detect outliers, which identified six outliers. After removal of these outliers, the Cochran’s Q statistic indicated no evidence of heterogeneity (P = 0.31 in IVW and P = 0.36 in MR-Egger), while the MR-Egger intercept remained non-significant (P = 0.90), indicating no evidence of directional pleiotropy. The two-sample MR without outliers yielded evidence for a significant causal association between pineal gland volume and daytime napping (IVW beta = −0.014, P_FDR_= 0.006; weighted median method beta = −0.018, P_FDR_= 0.026). All five MR methods showed consistent estimates after removal of the outliers (**Figure 4a,b),** supporting the robustness of this finding. In reverse MR, we also found a suggestive causal relationship between daytime napping and pineal gland volume (**Supplementary Table 21**). However, this association did not survive correction for multiple comparisons (IVW beta = −0.219, P_FDR_= 0.20) (**Figure 4c)**.

**Figure 4:**
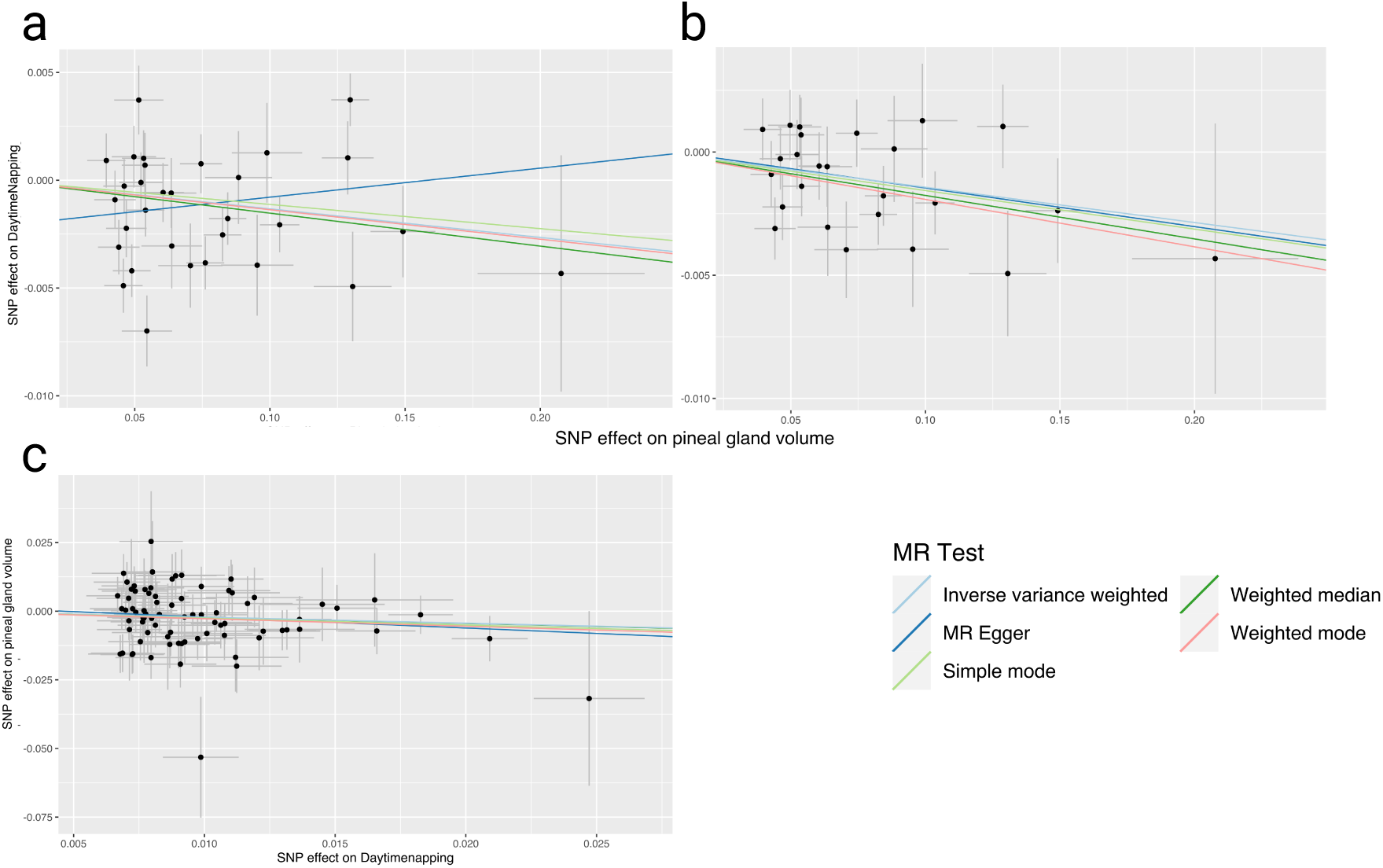
MR analyses for daytime napping and pineal gland volume. Scatter plots illustrating SNP effects on daytime napping versus pineal gland volume in forward MR analysis across five different MR methods: **a.** with and **b.** without removing outliers. **c.** Scatter plots representing SNP effects on pineal gland volume versus daytime napping in reverse MR analysis across five different MR methods. **Abbreviation:** GWAS, genome-wide association study; MR, mendelian randomization; SNP, single nucleotide polymorphism.

We found no evidence for causal associations between pineal gland volume and other sleep-related or neurological traits/disorders (**Supplementary Tables 20** & **21**).

## Discussion

Leveraging comprehensive genetic and brain imaging data from two large independent population-based studies, we performed the first GWAS of pineal gland volume as a proxy of melatonin production. Results across both cohorts were highly consistent, resulting in the detection of 34 genomic loci in the meta-GWAS. Notably, the SNP heritability of pineal gland volume was 19%, indicating that, like other brain regions^50^, the size of this key neuroendocrine structure is under substantial genetic control. Gene mapping and functional annotation highlighted multiple pathways involved in the regulation of a broad range of functions, especially sleep, metabolism, neuronal development and function. Furthermore, we observed genetic correlations between pineal gland volume and specific brain imaging-derived endophenotypes, especially volumes of structures located in the posterior cranial fossa and CSF, indicating a shared genetic architecture. Importantly, two-sample MR provided evidence for a causal relationship between pineal gland volume and daytime napping, suggesting that larger pineal gland volume is causally linked to reduced daytime napping.

We discovered 21 genes that were robustly associated with pineal gland volume through various gene mapping approaches. These included 15 genes (12 protein-coding and three RNA genes) located within the identified genomic loci in both the discovery and replication cohort, and six genes consistently identified across four complementary mapping methods (i.e., MAGMA, and positional, eQTL, and chromatin interaction mapping). Among these 15 genes, *MIR206* (tagged by *rs1892277),* which encodes the microRNA miR-206, emerged as the strongest candidate. As a post-transcriptional regulator, miR-206 influences the expression of key proteins critically involved in neuronal development, differentiation and function, including brain-derived neurotrophic factor, histone deacetylase 4 and the transcription factor JunD that protects cells against senescence and apoptosis^58–60^. Interestingly, elevated levels of miR-206 have been associated with cognitive decline, mild cognitive impairment and Alzheimer’s disease dementia^61^. Although the association between melatonin and miR-206 has not been assessed before, our findings suggest that part of melatonin’s neuroprotective effects may be mediated through its influence on miR-206 expression, warranting further experimental validation.

Many of the other genes identified to be associated with pineal gland volume also contribute to neuronal structure and function. For instance, *ZIC4* encodes a zinc finger protein involved in neurodevelopment^62^, and was also associated with the volumes of other brain structures in a previous GWAS study^50^. Similarly, *COL25A1* is also involved in neurodevelopment^63^, while *MST1R, RBM6, DCDC1, ETV3L, C12orf42, THADA, IMMP1L, ALX4, TEX41* and *DIRC3* have been associated with different brain volumetric measurements^50,64–67^. These genes may thus primarily exert their influence through effects on pineal gland development. Additionally, *CAMKV* encodes the CaM kinase-like vesicle-associated protein, which is involved in the regulation of synaptic transmission^68^, while *NCKAP1* and *RIMS2* also play roles in synaptic plasticity and function^69,70^. These latter genes are particularly relevant to pineal gland activity, which is regulated through a polysynaptic pathway originating in the melanopsin-containing retinal ganglion cells and reaching the pineal gland via the hypothalamic suprachiasmatic and paraventricular nuclei, and the superior cervical ganglion^71^. Importantly, multiple other genes associated with pineal gland volume, including *CAMKV*, *ETV3L, IGF1R, THADA, COL25A1, BEND7* and *ALX4*, have previously been linked to neurodegenerative diseases^72–76^, implicating them as promising candidate genes for disentangling the mechanisms through which melatonin affects neurodegeneration.

A large proportion of the genes associated with pineal gland volume have previously also been linked to sleep-related and metabolic traits. Specifically, nine genes, including *MST1R, DCDC1, RBM6, DNAJC24, CAMKV, ZIC4, BEND7, IGF1R* and *IMMP1L* were previously associated with insomnia^77,78^. Among these, *CKMKV* has been linked to chronotype as well^79^, while *RBM6* was related to both chronotype^79^ and sleep duration^80^. Similarly, *THADA* has been associated with obstructive sleep apnea^81^, whereas *C12orf42* has been linked to daytime napping^82^. Moreover, polymorphisms in *IGF1R, EDN2, COL25A1, RIMS2, BEND7, ALX4, DIRC3, THADA, C12orf42, MST1R, RBM6, DNAJC24, ZIC4, TEX41* and *CAMKV* have been linked to various metabolic traits, including height^83^, body mass index^84–89^, obesity and glucose and insulin signaling^90^. Importantly, in two-sample MR, we found evidence for a causal connection between larger pineal gland volume and reduced daytime napping. This finding extends prior reports on the association of daytime napping with disturbed melatonin secretion^91,92^. Collectively, these findings indicate that the genetic architecture of pineal gland volume closely reflects biological processes regulating sleep and metabolism, paralleling the crucial role of melatonin in regulating sleep-wake cycles and metabolism. Future studies should focus on how the expression of these genes affects melatonin secretion, and vice versa.

We also observed significant genetic correlations between pineal gland volume and several brain imaging-derived endophenotypes, notably the volumes of the isthmus of the cingulate gyrus, the cerebellum vermis lobules I-V and CSF. These associations may partly reflect the shared development and close anatomical proximity of the pineal gland and these structures. The pineal gland develops as an evagination of the dorsal diencephalical neuroepithelum, and is located just below the isthmus cinguli and above the vermis cerebelli^93^. Interestingly, pineal gland volume has previously also been associated with mood disorders^94^, which in turn were also related to the isthmus of the cingulate^95^, a critical component of the limbic system, as well as the cerebellum vermis lobules I-V^96^. Thus, also shared genetic mechanisms underpinning the morphology and/or function of these structures and mood disorders may be involved. Furthermore, a previous study reported that larger CSF volume was significantly associated with improved sleep^97^. Although the exact mechanisms underlying this relation remains unclear, our findings raise the possibility of shared genetic factors impacting both melatonin production and CSF circulation. Thus, genetic correlations between pineal gland volume and other brain structures point towards common genetic factors influencing melatonin production, sleep/circadian rhythm regulation, and CSF circulation.

Our study has several potential limitations. First, the sample size of our replication GWAS was relatively small. Nevertheless, the effect estimates were highly consistent between the discovery and replication GWAS. Indeed, we were able to replicate about two-thirds of the genetic associations identified in the discovery GWAS, firmly supporting the robustness of our findings. Second, only people from European ancestry were included in our analyses due the fact that they represented the overwhelming majority of participants enrolled in both cohorts. Therefore, further GWAS studies in other ethnic populations are warranted for extending the generalizability of our findings.

In conclusion, our findings elucidate the genetic architecture of the pineal gland, and identify numerous novel candidate genes and molecular pathways that may underlie the effects of melatonin on a broad range of functions, especially sleep/circadian rhythm, metabolism, neuronal development and brain health.

## Contributors

PX, MAI, MMBB, and NAA conceptualized the project. The first draft of the manuscript was written by PX, MAI and NAA. Data were analyzed by PX, MAI, and NAA. DR, SE, MR and MMBB provided technical, statistical and methodological advice. All authors provided critical feedback and contributed to the writing and revision of the final version of the manuscript.

## Acknowledgments

We would like to thank the Rhineland Study team for supporting the data acquisition and management. This work was supported by DZNE institutional funds, the Federal Ministry of Education and Research of Germany (FKZ: 031L0206, 01GQ1801, 01KX2230), the Helmholtz Association under the 2023 and 2024 InnovationsPool, the Deutsche Forschungsgemeinschaft (DFG, German Research Foundation, Project-ID 432325352), an Alzheimer’s Association Research Grant (Award Number: AARG-19-616534), the Chan Zuckerberg Initiative (Project FastSurfer, Grant Number: EOSS5 2022-252594), the Helmholtz-AI project DeGen (ZT-I-PF-5-078), and NIH (R01 LM012719, R01 AG064027, R56 MH121426, and P41 EB030006). Peng Xu is supported by a scholarship from China Scholarship Council (Number:202108310025), and N. Ahmad Aziz is supported by a European Research Council Starting Grant (Number: 101041677). Part of this research has been conducted using the UK Biobank Resource under application number 82056.

## Data availability

The Rhineland Study’s dataset is not publicly available because of data protection regulations. Access to data can be provided to scientists in accordance with the Rhineland Study’s Data Use and Access Policy. Requests for further information or to access the Rhineland Study’s dataset should be directed to. All individual level data from the UK Biobank Imaging Study used in the current manuscript are available through the UK Biobank Resource (https://www.ukbiobank.ac.uk). This research has been conducted using the UK Biobank Resource under application number 82056. We used publicly available GWAS summary statistics for chronotype (https://journals.sagepub.com/doi/full/10.1177/0748730420935328), daytime napping and sleep duration (https://www.nature.com/articles/s41467-020-20585-3), cognition (https://www.nature.com/articles/s41588-018-0152-6), Alzheimer’s disease (https://www.nature.com/articles/s41588-022-01024-z), Parkinson’s disease (https://pmc.ncbi.nlm.nih.gov/articles/PMC8422160/), and brain imaging-derived endophenotypes (https://www.nature.com/articles/s41588-019-0516-6).

